# Search for trends of Covid-19 infection in India, China, Denmark, Brazil, France. Germany and the USA on the basis of power law scaling

**DOI:** 10.1101/2020.04.03.20052878

**Authors:** Srijit Bhattacharya, Alokkumar De, Md Moinul Islam

## Abstract

The corona virus (SARS-CoV-2) or Covid-19 pandemic is growing alarmingly throughout the whole world. Using the power law scaling we analyze the data of different countries and three states of India up to 1st April, 2020 and explain in terms of power law exponent. We find significant reduction in growth of infections in China and Denmark (γ reduced from approximately 2.18 to 0.05 and 11.41 to 6.95, respectively). Very slow reduction is also seen in Brazil and Germany (γ reduced from approximately 6 to 4 and 11 to 7, respectively). Infection in India is growing (γ=9.23) though lesser in number than that in the USA (highest γ of 16 approximately, studied so far), Italy and a few other countries. Among three Indian states the growth in West Bengal (γ=0.64) is much slower than other states like Maharashtra and Kerala (γ=3.23 and 3.32, respectively). Some future predictions, though not rigid, has also been incorporated in our analysis. The earlier lock-down and stricter measures from the Governments concerned are being thought to be the only possible solutions, in the present situation, to fight against this virus.

## Introduction

An epidemic implies an increase, often sudden, in the number of cases of a disease above the normally expected level in the population of an area. Although outbreak carries the same meaning of epidemic, but generally is used for more limited geographical area. However, pandemic is completely different from both the terms on the basis of the outbreak area and affected people as defined in [1]. It generally spreads over several countries or even continents impacting very large number of people. Pandemic is often caused by novel or new virus or new strain of virus [1]. The disease spread out from the novel virus is generally dangerous owing to the lack of human immunity. The Spanish flu in 1918-19 and H1N1 in 2009 are concrete examples of some of the deadliest pandemics. The former killed almost 40 million people globally infecting a quarter of the then global population. 2009 H1N1 flu was another example that infected about 163258 people (confirmed case), though the mortality rate was small (only 18036 was confirmed by World Health Organization) [2, 3, 4, 5].

Presently, human race is in a precarious condition due to the infection of a novel corona virus or, i.e., COVID-19. Identification of the virus was done in Wuhan of China in December, 2019 and officially named as “severe acute respiratory syndrome coronavirus 2 (SARS-CoV-2)” by the International Committee on Taxonomy of Viruses (ICTV). Since then the pandemic spread globally and presently total confirmed cases Worldwide is more than 905209 till 1st April, 2020 11:59 pm. Out of the infected persons, 45371 (5%) have been killed and nearly 190710 (∼21.1%) have been recovered as of 31st March, 2020 11:59 pm. Initially the outbreak was maximum in China. But now the disease is spread in different countries. The USA is highest in terms of confirmed cases. Italy has registered highest death of 13155 till now as per the date and time mentioned above. The recovery is maximum (∼76405) in China at the same time and date [6, 7].

Under this condition, the Governments of different countries have come forward with different ideas to save the citizens from the peril of the virus. However, all the Governments have uniformly accepted that due to the non-existence of vaccines or globally accepted medications at present, lock-down and social distancing could be the solutions to save the countries from uncontrollable infections. Some countries e.g Denmark, Norway and South Korea have significantly reduced the rate of infections by the interventions like emergency lock-downs, and social distancing much earlier than other countries [8]. Under these circumstances it is important to understand the characteristics of the COVID-19 infections, the rapidity of its spread and recovering from the ailments concerned.

Simple mathematical models may be used at present to understand grossly the trend of the already observed infections, as well as to predict its future behavior. At the very moment, there is tremendous need of predictive models or formulae to understand the extent of global spread in the near future so that effective and efficient plans could be adopted in different sectors of medical field and Government policies. Recently, two approaches have been used to estimate the infection rate. The SIR (susceptible infected recovered) model for the spread of infectious disease primarily has been used to predict the number of confirmed COVID-19 cases [9, 10]. Besides, logistic models [11, 12], time series modeling [13] etc are also used for the same. Another approach is to use some simple models like power law, exponential and parabolic (t^2^) scaling of growth etc. Among these, power law model is stated to be free from some errors as seen in exponential growth and parabolic growth models [8, 14,15], and it is represented by:

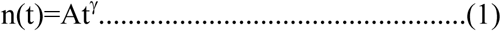

where, γ is the power law exponent and A is constant. The estimation of the exponent (γ) in the power law is done by fitting the available data on the total number of infected persons as a function of time. The value of γ signals the rapidity of the spread of disease in a country or region. Sengar [8] found that the number of confirmed cases had reached a plateau or saturation (new infection reported becomes zero) in China. Except China, severe reduction in new confirmed events is found only in a few other countries such as Denmark, Norway and South Korea but not collapsed to zero so far as the time span is concerned, Senger performed the calculations up to 24th March, 2020 to see whether there was significant reduction in the exponent (γ) during the Government intervention and if the emergence of saturation in the number of infection could be imminent [8]. In the present work, we have done the same type of calculations for several countries but extending the date up to 1st April, 2020 to examine the value of the exponent and to find out the effect of Government intervention in the prevention of the pandemic. Apart from India as a whole, we have also performed calculations for its three states, viz. Kerala, Maharashtra and West Bengal, separately. Short term future prediction has also been incorporated in the present work.

## Materials and Method

The data for all the countries studied, except India, were extracted from the JHU-CSSE 2020 open data source for COVID-19. JHU-CSSE is an endeavour from John Hopkins University [7]. We have used the data available in the above domain to find the cumulative total number of infected persons as a function of elapsed time. The data for different countries like China, France, Germany, the USA, Brazil, Denmark etc have been extracted and plotted in ordinary as well as in log-log scale. Similarly, the data have also been plotted for India along with its three states-Kerala, Maharashtra and West Bengal, separately. The data of the three states are available from the Ministry of Health and Family Welfare, Govt. of India Website [16]. The power law fit is done for each countries in the log-log scale and the least square best fit value of the exponent is estimated from the straight linear portion of the plot.

## Results and Discussions

Results have been displayed in the following figures (figs.1-9). The data is plotted in ordinary scale and also in log-log scale for different countries. Fig 1 is the cumulative total number of infected persons plotted against total number of days elapsed for China. Fig. 2 depicts the same data but represented in log-log scale. Similar plots in normal and log-log scales concerned are shown in figs. 3-4, 5-6 & 7-9 for Germany, India and three Indian states, respectively. The plots corresponding to other countries studied here has not been shown in the paper, though calculations have been performed. In all the plots, the cumulative total number of days elapsed scale starts from the respective first day of infection (it varies from 22 January, 2020 to 31st January, 2020 for different countries). The number of confirmed infected persons in China is very large (548 persons) at the very first day. Thus the graph of China Covid-19 infection comes much different from that of other countries.

**Fig 1:**
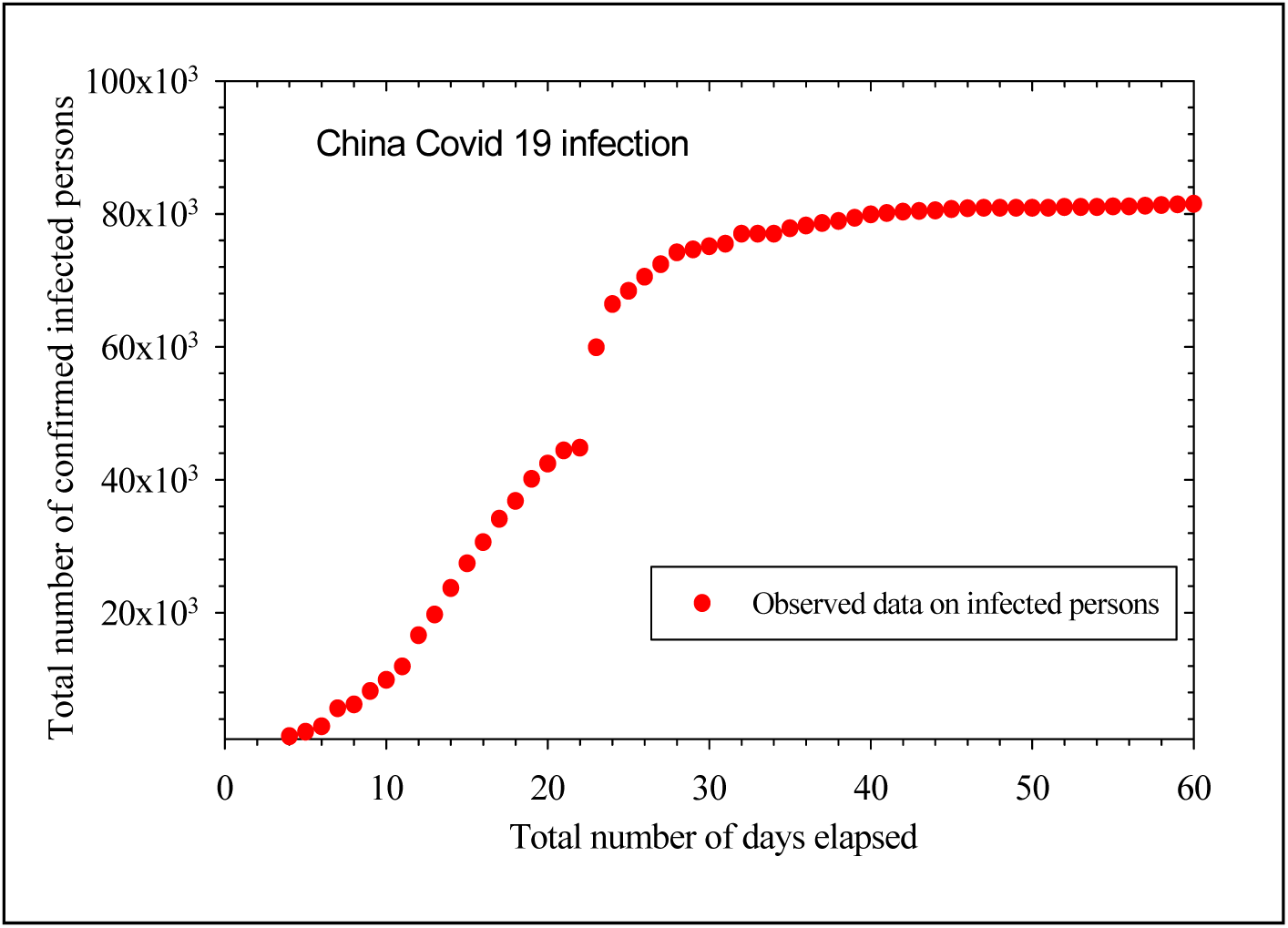
Covid-19 infection in China, plotting being done in normal scales. (starting date 22nd January, 2020)

**Fig 2:**
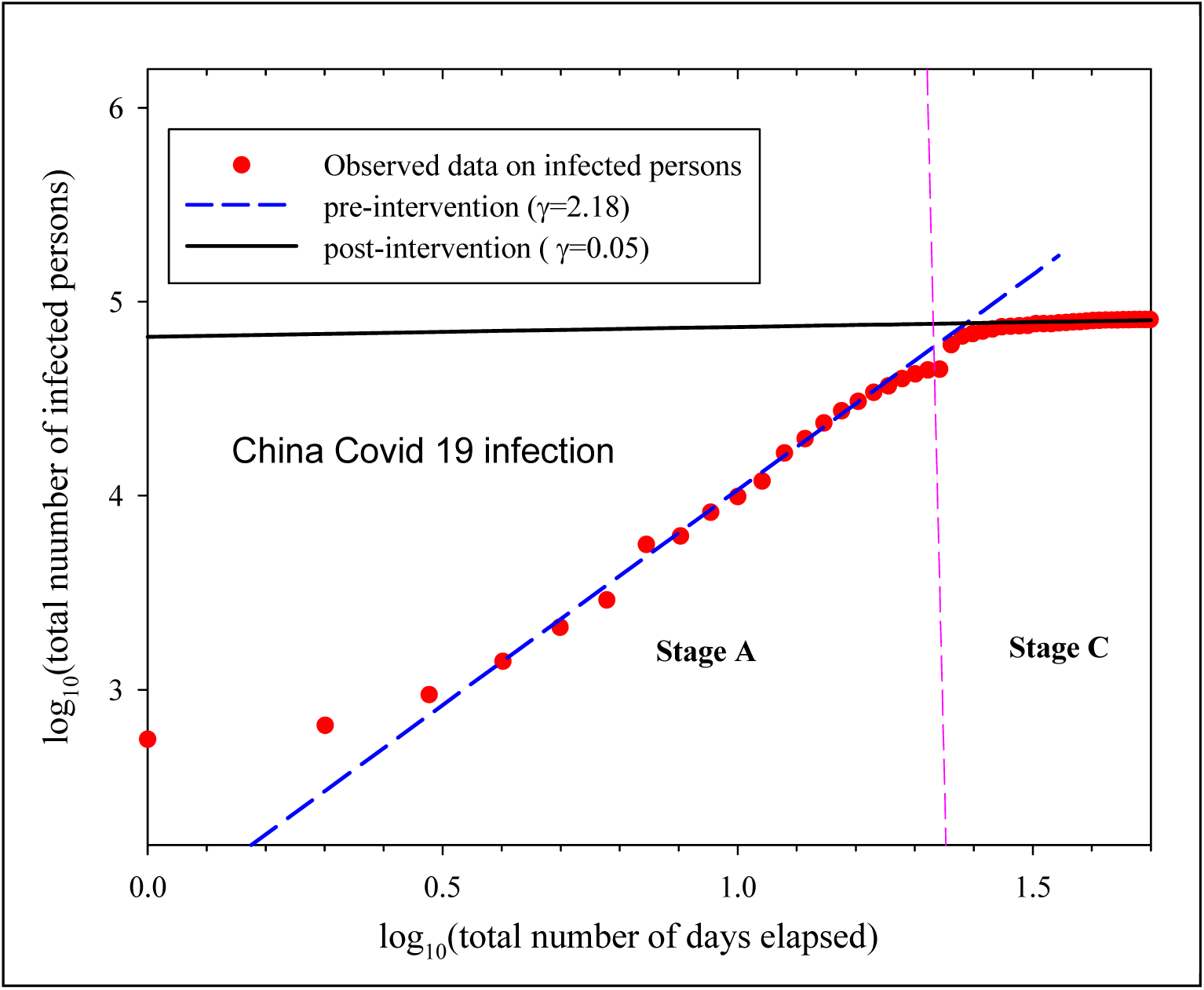
: Covid-19 infection in China, plotting being done in log-log scale. The two straight lines (blue dashed and black solid) are the yields of linear regression. The dashed straight line gives the value of exponent at stage A, while saturation stage or stage C is also shown. The two stages are divided by pink long dashed vertical boundary. The corresponding exponents (γ) are mentioned in the legend.

**Fig 3:**
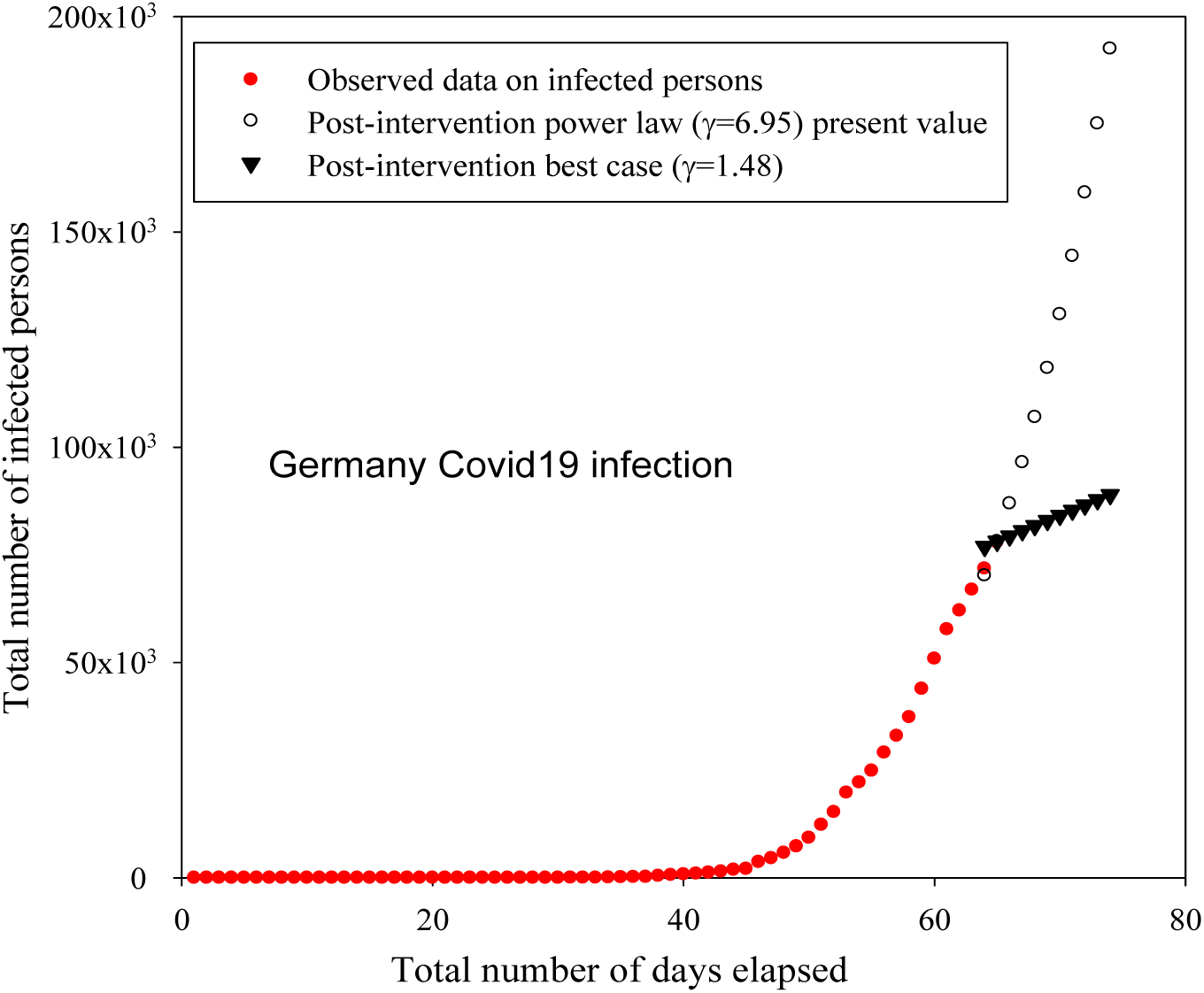
Covid-19 infection in Germany, plotting being done in normal scales. (starting date 27nd January, 2020). The prediction of numbers for 9 days next to 1st April, 2020 is also shown by black open circles (assuming γ_post_ =6.95, as estimated from fitting its data) for the worst case and black filled inverted triangles for the best case (assuming γ_post_ =1.48, as that of Denmark).

As shown in [8], manifestation of three different stages could be seen from the log-log plot of the cumulative total number of infected persons against the total number of days elapsed.

A. the growth stage-: in this stage the total number of infected persons can be fitted by power law.(eq.1). We fit this region by eq. 1 and extract the value of the exponent (γ).
B. the slow down stage:-here the effect of Government measures can be visible through the reduction of the exponent (γ).
C. the saturation stage:- no new infections are reported and the curve tends to a plateau. The value of γ becomes very close to zero.

In majority of the countries the stage A can only be seen, while no slow down stage is found. In only a handful of countries, e.g Denmark, South Korea and Norway the stages A and B are prominently visible without any sign of stage C [8]. Only China is showing the stages A and C. The stage C or saturation comes just after the end of the growth stage. For majority of the countries the initial growth of the disease is very rapid and as the Government intervention comes into effect the log-log plot moves to the power law region or stage A. We have excluded a few points at the very beginning during this rapid growth in the straight line fitting in our work. The value of the exponent at stage A is termed as pre-intervention exponent (γ_pre_) and that at stage B or C is termed as post-intervention exponent (γ_post_). Till now the co-existence of stages B and C have not been found for any countries we have analyzed.

From fig. 2 the data of China is found in agreement with the earlier work [8] having best fit pre-intervention exponent γ_pre_=2.18±0.04 and post-intervention exponent γ_post_=0.05±0.018. The reduction of γ is significant that may indicate the faster and stricter Government measures and adherence of rules by the citizens. Similarly, we estimated the exponent γ for Germany, France, Brazil, Denmark and India. Our intention is to check how much changes could be found in the data within a few days of the earlier published work and also to investigate whether saturation stage comes into effect in any countries other than China.

Earlier it was found that there was rapid reduction in the rate of number of infected persons only in China, Denmark, South Korea and Norway. However, except China, the perfect saturation stage of infection could not be seen in the rest of the countries corroborated by the higher values of post-intervention γ in comparison to that of China. The γ_post_ was found nearly zero in China. For Denmark, we found γ_post_ as 1.48 similar to the earlier work. Figs 3 & 4 denote the total number of patients for Germany as a function of total number of days elapsed in ordinary and log-log scales, respectively. In our work we have also found that in Germany (fig. 4) and Brazil (not shown in figure) the saturation stage has just been started. The γ_post_ =6.95±0.08 is lesser than γ_pre_=11.41±0.14 in Germany. But the reduction of γ_post_ is not as severe as that of China or even Denmark. Thus it is not confirmed whether true saturation region is actually started there or it is simply a daily fluctuation of the data. The prediction of the total number of infections is shown in fig. 3 taking γ_post_=6.95 and also by taking γ_post_ same as that of Denmark for the best case scenario. Table 1 shows the values of exponent for different countries and comparison with the earlier results.

**Table 1:** The pre and post-intervention γ for different countries including India along with three Indian states. (Source:[6] and [13]).

|  | Our work upto 1st April, 2020 |  | Earlier work upto 24 March, 2020 |  | Number of infection as on 1st April, 2020 12 midnight IST |
| --- | --- | --- | --- | --- | --- |
| Country | $\gamma_{\text{pre-intervention}}$ | $\gamma_{\text{post-intervention}}$ | $\gamma_{\text{pre-intervention}}$ | $\gamma_{\text{post-intervention}}$ | |
| China | 2.18 | 0.05 | 2.22 | 0.024 | 82300 |
| Germany | 11.41 | 6.95 | 11.58 | ----- | 66900 |
| Brazil | 6.20 | 4.17 | 6.39 | ----- | 4600 |
| The USA | 15.91 | ----- | 16.59 | ----- |  |
| Denmark | 6.82 | 1.48 | 6.82 | 1.48 | 2800 |
| France | 10.81 | ----- | 10.14 | ----- | 57700 |
| India | 9.23 | ----- | 9.76 | ----- | 1834 |
| Kerala | 3.32 | ----- | ----- | ----- | 265 |
| Maharastra | 3.23 | ----- | ----- | ----- | 335 |
| West Bengal | 0.64 | ----- | ----- | ----- | 37 |

**Fig 4:**
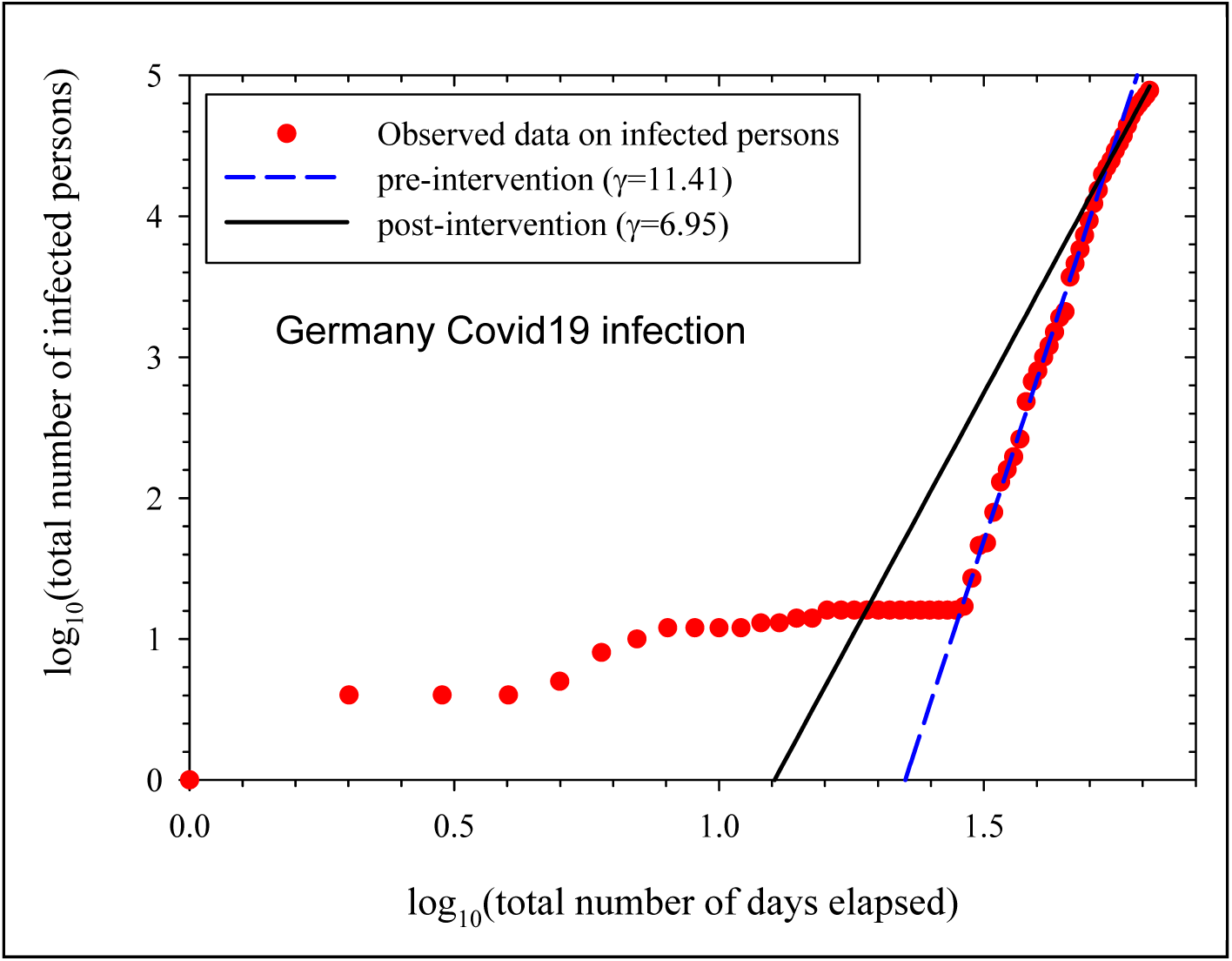
The observed total number of infected persons is plotted against total number of days elapsed for Germany in log-log scale and the best fit straight lines are shown. The reduction in the value of exponent is shown in the legend.

We also analyzed the data as observed in India but the appearance of stage B or C could not be found as on 1st April, 2020. However, the pre-intervention γ is found as 9.23±0.17, lesser than that found in the earlier work. Fig. 5 depicts the data on cumulative total number of infected persons as a function of total number of days elapsed and fig.6 gives the log-log plot of the confirmed infection data. However, one could predict the probable future situation of India by imposing, with immediate effects, the available post-intervention best (γ_post_=1.48) and worst γ-values (γ_post_=6.95) as available from the data of other countries. The result is shown in fig.5 & table 2.

**Table 2:** Our short term prediction of cumulative total number of infected persons in India.

| Date | With $\gamma=9.23$ (similar to the present trend ongoing in India) | With $\gamma=1.48$ best post-intervention case (similar to Denmark) | With $\gamma=6.95$ in worst post-intervention case (similar to Germany) |
| --- | --- | --- | --- |
| 2nd April, 2020 | 2109 | 1870 | 2072 |
| 5th April, 2020 | 3220 | 2002 | 2849 |
| 8th April, 2020 | 4824 | 2136 | 3862 |
| 14th April, 2020 | 10307 | 2412 | 6840 |

**Fig 5:**
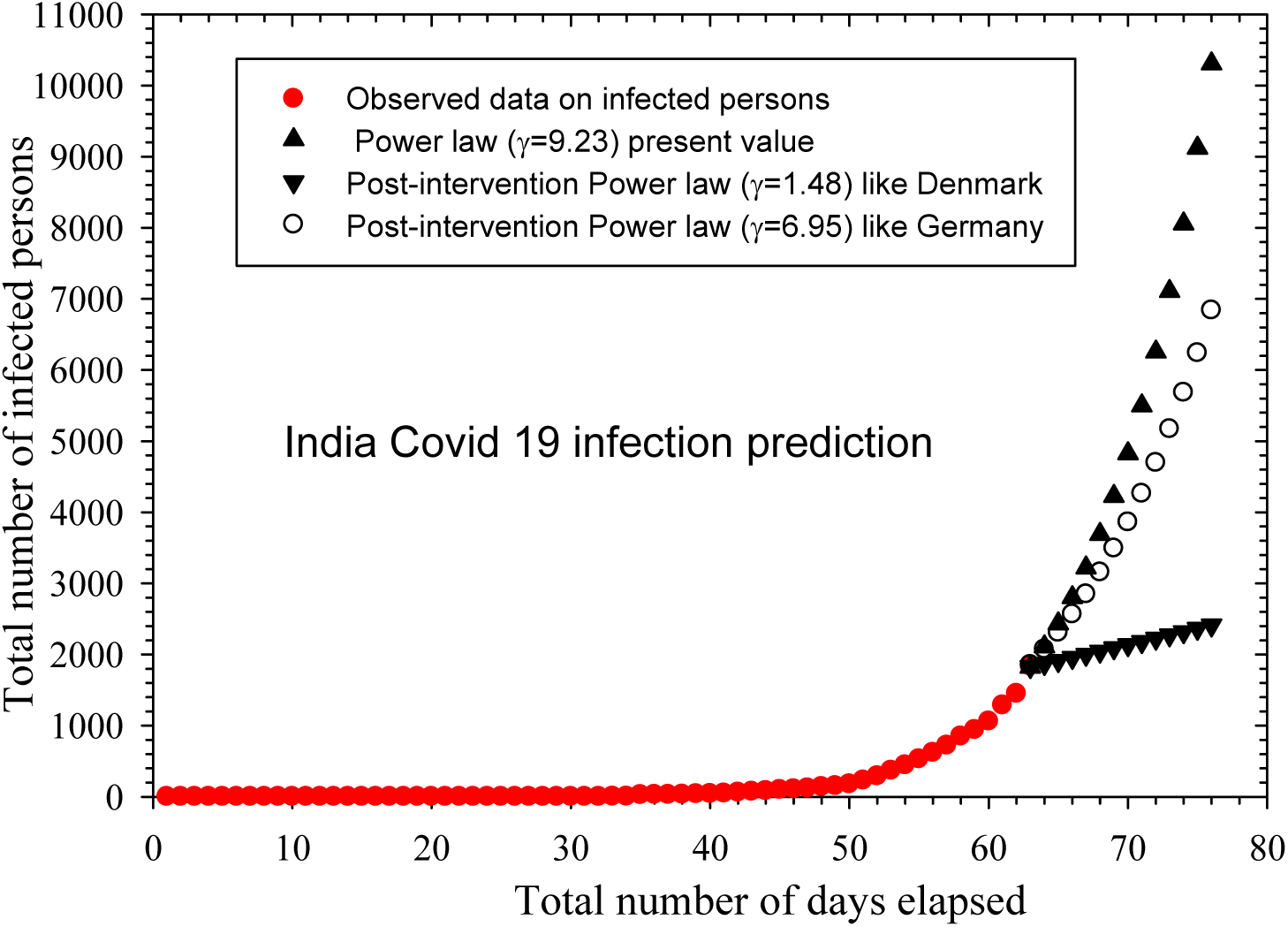
Covid-19 infection in India, plotting being done in normal scale. Short term predictions are also shown for three possible cases (black filled triangles, unfilled circle and filled inverted triangles) as shown in the legend. (Starting date 30th January, 2020).

Among the three states of India, the growth of infection in Kerala (Fig. 7) and Maharashtra (Fig. 8) is much higher than that in West Bengal (Fig. 9). However, post-intervention saturation effects could not be seen in any of these states. Table 1 shows the list of exponent values and number of infections in different countries and Indian states.

**Fig 6:**
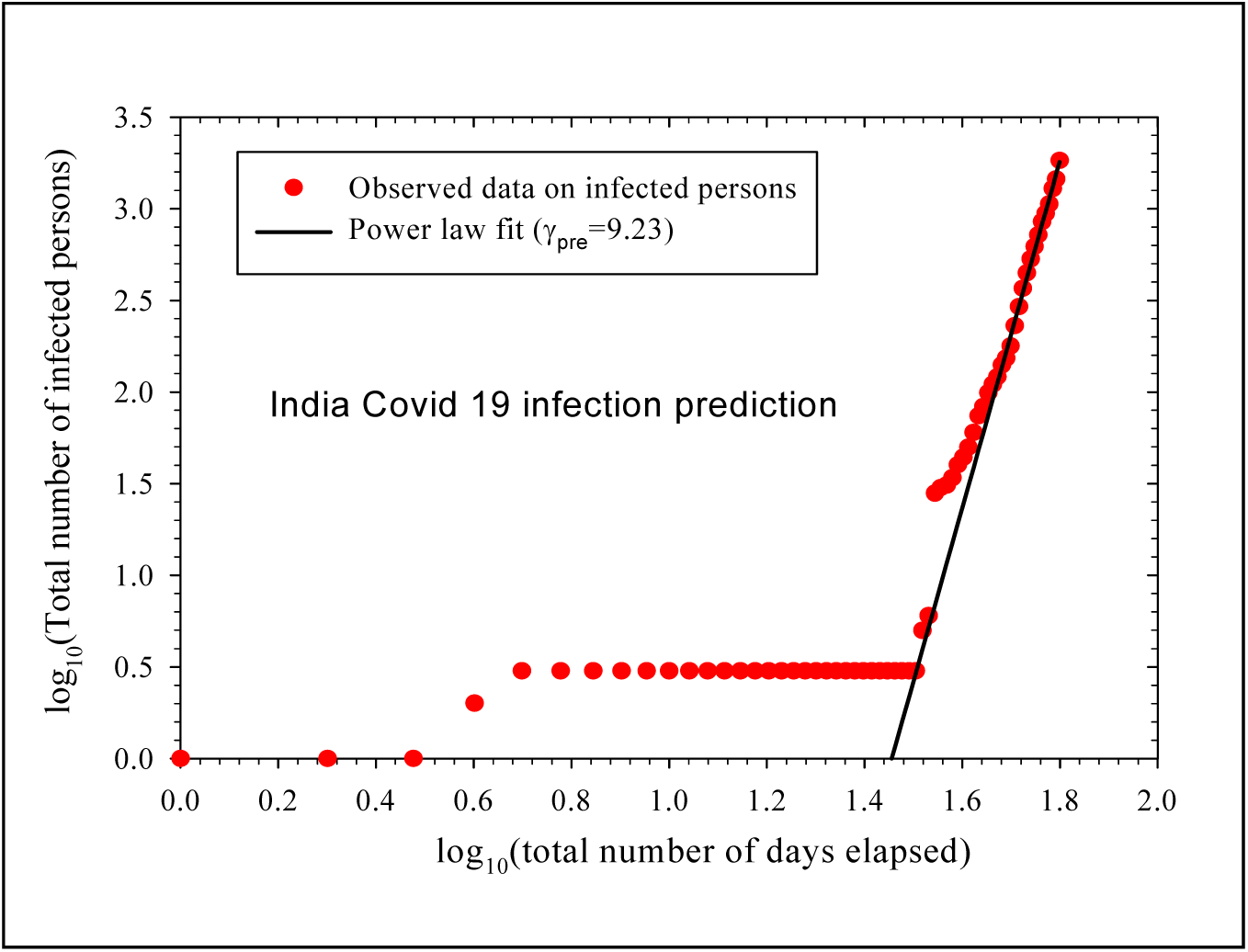
log-log plot of infection in Indian perspective along with best fit straight line to estimate γ.

**Fig 7:**
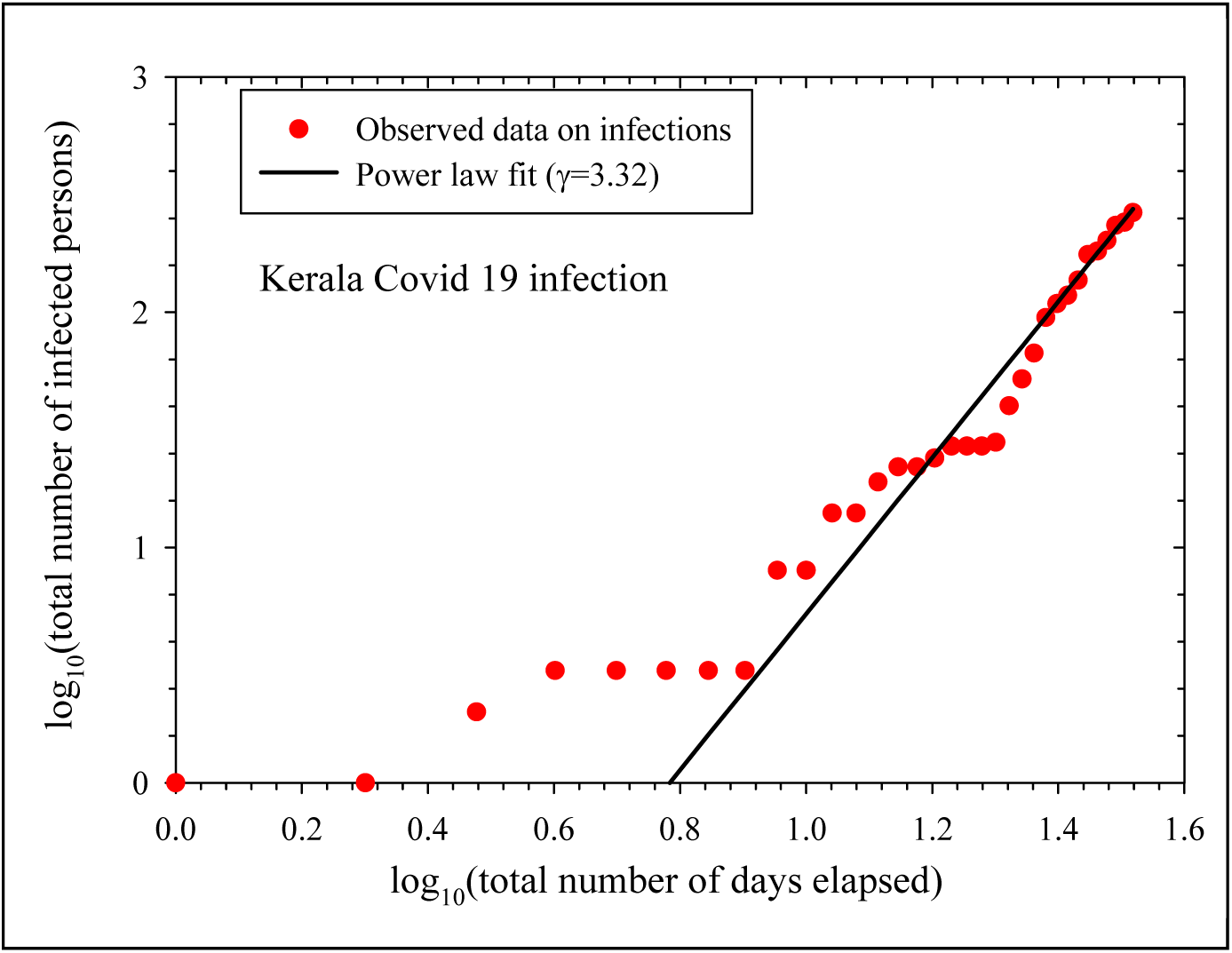
log-log plot of infection along with best fit straight line to estimate γ for the state Kerala (starting date 30 January, 2020).

**Fig 8:**
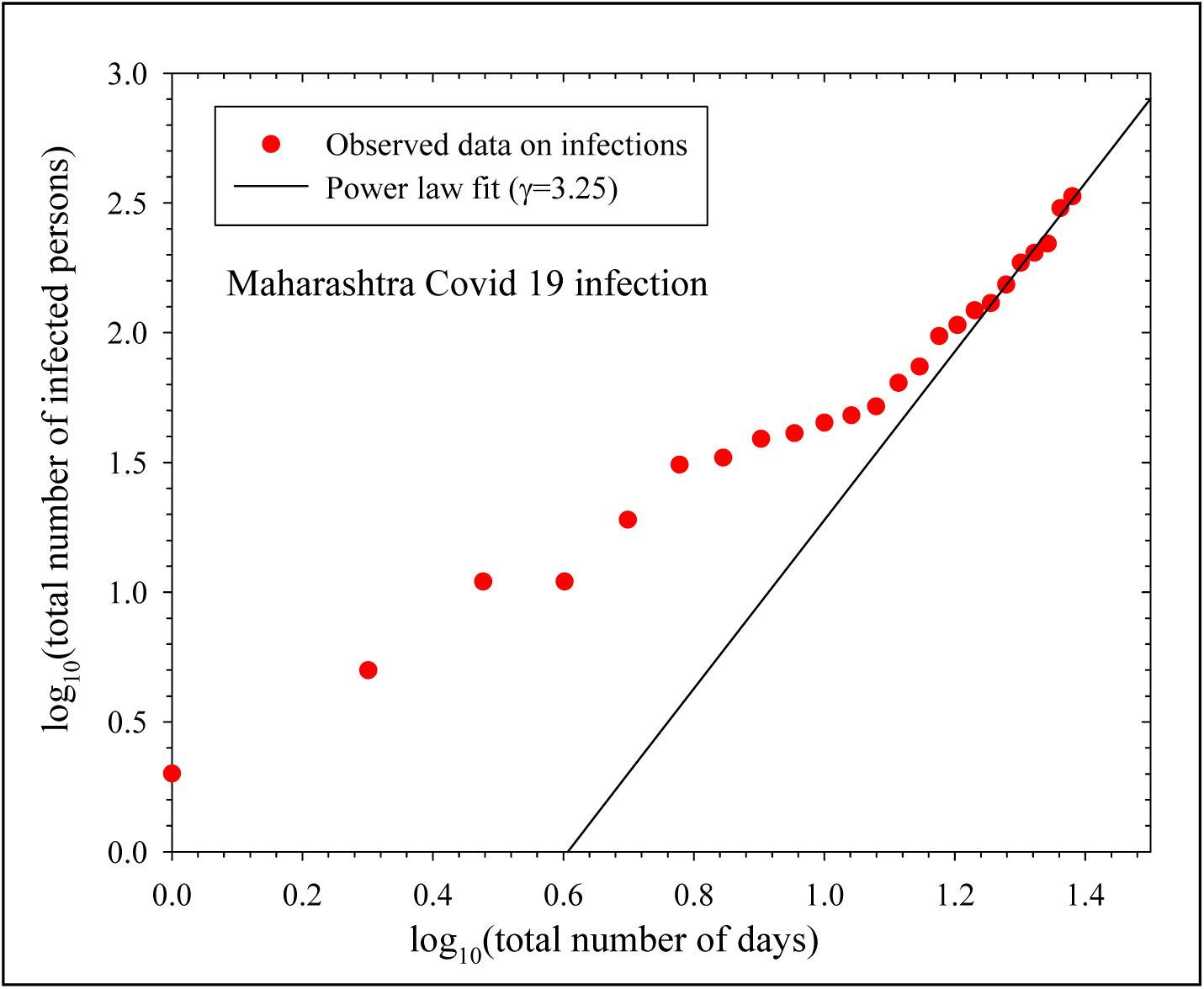
log-log plot of infection along with best fit straight line to estimate γ for the state Maharashtra (starting date 9th March, 2020).

**Fig 9:**
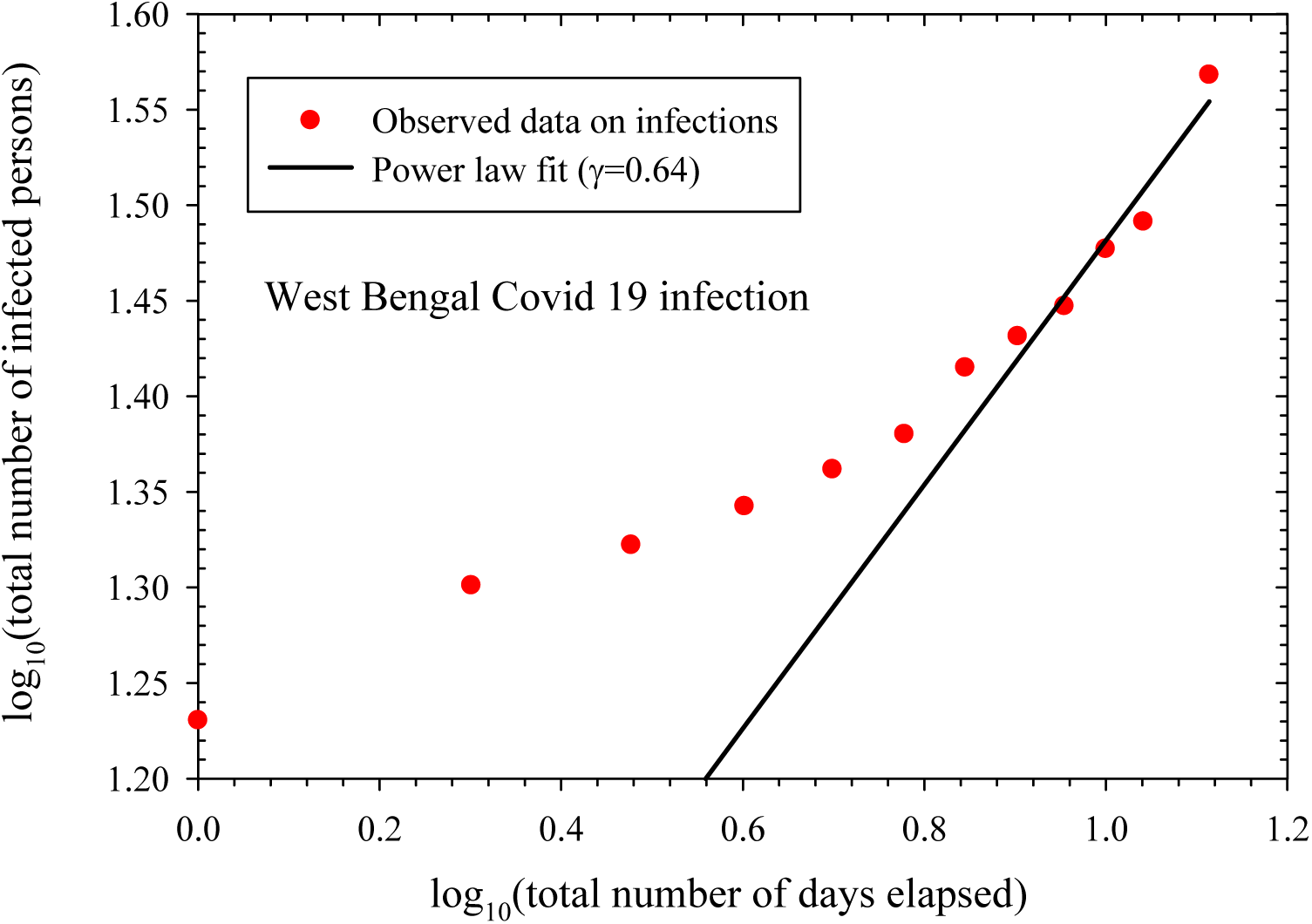
log-log plot of infection along with best fit straight line to estimate γ for the state West Bengal (Starting date 17th March, 2020).

We feel the reduction of exponent in the power law as well as the appearance of saturation region in the data depend on different factors such as the rapid intervention from the Government, adherence of rules by the citizens, facilities of treatment etc. The rate of infection in India is still increasing following power law. However, the extent of infection in terms of confirmed number of infected persons is lower in comparison to those of other countries such as the USA (highest γ=15.91), France or Italy, if we take into account the larger population of India. However, the citizens should also come forward actively and efficiently to help materialize successful implementation of Government policies by adhering the Government rules. However, one thing is certain, as commented in earlier work [8], that earlier and stricter lock-down aided by general public co-operation may reduce the infection rapidly as shown by China, South Korea and Denmark. The effect of lock-down in India may be visible in the next few days, affecting the value of the exponent of the power law scaling. Similar case is also seen for the states studied for the country India. West Bengal is one of the Indian States where lock-down started from as early as 14th March, 2020. It may be one of the reasons for the slower growth in infection in West Bengal in comparison to other states. In case of the USA, largest value of γ is really unexplained. But we have found a ray of hope. The USA data of last 5 days may be showing reduction in γ (from 15.91 to as small as 9), though the number of data points is very small to comment.

Our research work is still preliminary one. The predictions may vary for different countries according to social, economical, medical and ethical perspectives. Moreover, we found different data sources (covid19india.org, World Health Organization, John Hopkins University database, Worldometer etc) along with Indian Government Website. Since different data sources are showing variable daily data, we have strictly adhered to Indian Government website. The present work is nothing but some sincere endeavour to understand the trend of the ongoing situation. This might be helpful for the respective administrations to adopt adequate measure in fighting against the infection concerned. If we can foresee the trend of the infection, it will be easier to preplan to fight against the virus.

## Conclusions

We have studied the nature of gradual spread in cumulative total number of infected people in several countries like China, Denmark, Germany, Brazil, the USA along with India on the basis of power law scaling. The spread in the number of infections are also studied in the three states (Kerala, West Bengal and Maharashtra) of India separately. The common trend is the initial rapid growth stage of infection with time and then the reduction of rate due to the intervention of the respective Governments. In China, the actual saturation in the new events has been found. The pre and post saturation exponents γ are found similar to earlier work. Denmark, Brazil and Germany show reduced post saturation exponent γ indicating the appearance of stage B. The German and Brazilian data may have showed the post-intervention effect due to Governmental policies with γ_pre_ and γ_post_ being different. The data on infected persons show that India still has not reached stage B. The stricter Government interventions may be visible in the data later. We have predicted the short term number of confirmed infected patients in best case (γ_post_ same as that of Denmark) and in worst case scenarios (γ_post_ same as of Germany) for India along with the trend due to present value of the exponent. Similar prediction has also been done for Germany. The German data set is interesting as it is probably entering in the stage B or slow down stage. Among the three Indian states, the rate of infection is found very small in West Bengal in comparison to that in Maharashtra and Kerala. This analysis could be strengthened by incorporating more data in the future.

## Data Availability

The data is taken from open source data repositories.

## Acknowledgement

The authors reports no funding related to this research and have no conflicting financial interests. One of the authors (S.B) acknowledges Mr. Sridip Bhattacharya for his help in finding the data sources and for productive discussions.

## References

[1] Principles of Epidemiology in Public Health Practice, Third Edition An Introduction to Applied Epidemiology and Biostatistics. Centre for Disease Control and Prevention. The United States..

[2] https://www.who.int/emergencies/diseases/novel-coronavirus-2019;

[3] P. Spreeuwenberg; et al. (1 December 2018). “Reassessing the Global Mortality Burden of the 1918 Influenza Pandemic”. American Journal of Epidemiology. 187 (12): 2561–2567. doi:10.1093/aje/kwy191.

[4] Pandemic (H1N1) 2009 – update 100”. World Health Organization (WHO). 14 May 2010.

[5] Robert Roos. “CDC estimate of global H1N1 pandemic deaths: 284,000”. CIDRAP, June 27, 2012.

[6] WHO. Coronavirus covid-19 global cases by the center for systems science and engineering (csse) at johns hopkins university (jhu). URL https://www.arcgis.com/apps/opsdashboard/index.html#/bda7594740fd40299423467b48e9ecf6. Accessed on 2020-03-25 10:30 CET.

[7] JHU CSSE. Novel coronavirus (covid-19) cases. URL https://github.com/CSSEGISandData/COVID-19. Accessed on 2020-03-24 11:00 CET.

[8] H. M. Sengar. Short-term predictions of country-specific Covid-19 infection rates based on power law scaling exponents. 2003.11997v1 [physics.soc-ph] 26 Mar 2020.

[9] R. Singh and R. Adhikari. Age-structured impact of social distancing on the COVID-19 epidemic in India. 2003.12055v1 [q-bio.PE] 26 Mar 2020.

[10] Franz-Josef Schmitt. A simplified model for expected development of the sars-cov-2 (corona) spread in germany and us after social distancing. https://arxiv.org/abs/2003.10891, 2020;

[11] Slav W Hermanowicz. Forecasting the wuhan coronavirus (2019-ncov) epidemics using a simple (simplistic) model - update (feb. 8, 2020). medRxiv, 2020. doi:10.1101/2020.02.04.20020461. URL https://www.medrxiv.org/content/early/2020/02/10/2020.02.04.20020461;

[12] K. Roosa, Y. Lee, R. Luo, A. Kirpich, R. Rothenberg, J.M. Hyman, P. Yan, and G. Chowell. Real-time forecasts of the covid-19 epidemic in china from February 5th to February 24th, 2020. Infectious Disease Modelling, 5:256–263, 2020. ISSN 2468-0427. doi: https://doi.org/10.1016/j.idm.2020.02.002. URL http://www.sciencedirect.com/science/article/pii/S2468042720300051.

[13] Soudeep Deb and Manidipa Majumdar. A time series method to analyze incidence pattern and estimate reproduction number of covid-19. https://arxiv.org/abs/2003.10655, 2020.;

[14] Duncan J. Watts and Steven H. Strogatz. Collective dynamics of ‘small-world’ networks. Nature, 393(6684):440–442, 1998. ISSN 1476-4687. doi:10.1038/30918. URL https://doi.org/10.1038/30918.

[15] D.J. Watts. Small worlds: the dynamics of networks between order and randomness, Vol. 9. Princeton University Press, 2004.

[16] https://www.mohfw.gov.in/ https://en.wikipedia.org/wiki/2020_coronavirus_pandemic_in_India; https://en.wikipedia.org/wiki/2020_coronavirus_pandemic_in_Kerala; https://en.wikipedia.org/wiki/2020_coronavirus_pandemic_in_Maharastra.; https://en.wikipedia.org/wiki/2020_coronavirus_pandemic_in_West_Bengal;

